# Taiwan Biobank: a rich biomedical research database of the Taiwanese population

**DOI:** 10.1101/2021.12.21.21268159

**Authors:** Yen-Chen Anne Feng, Chia-Yen Chen, Tzu-Ting Chen, Po-Hsiu Kuo, Yi-Hsiang Hsu, Hwai-I Yang, Wei J. Chen, Chen-Yang Shen, Tian Ge, Hailiang Huang, Yen-Feng Lin

**Author notes:** **Correspondence:** Yen-Chen Anne Feng, Yen-Feng Lin.

## Abstract

The Taiwan Biobank (TWB) is an ongoing prospective study of over 150,000 individuals aged 30-70 recruited from across Taiwan beginning in 2012. A comprehensive list of phenotypes was collected for each consented participant at recruitment and follow-up visits through structured interviews and physical measurements. Biomarkers and genetic data were also generated for all participants from blood and urine samples. We present here an overview of the genetic data quality, population structure, and familial relationship within TWB, which consists of predominantly Han Chinese-ancestry individuals, and highlight important attributes and genetic findings thus far from the biobank. A linkage to Taiwan’s National Health Insurance database of >25 years and other health-related registries is underway that will enrich the phenotypic spectrum of TWB and enable deep and longitudinal genetic investigations. TWB provides one of the largest biobank resources for biomedical and public health research in East Asia that will contribute to our understanding of the genetic basis of human health and disease in global populations through collaborative studies with other biobanks.

## Introduction

Situated in East Asia, Taiwan is an island of 36,000 km^2^ comprising a population of 23 million people with a well-established public health infrastructure. To advance epidemiological and biomedical research, Taiwan launched its own biobank, the Taiwan Biobank (TWB), in 2012, which prospectively collected a wide variety of lifestyle behaviors, environmental risk factors, and family history of common, complex diseases in the general Taiwanese population with genetic information also measured in the biobank participants. One powerful aspect of TWB is the ability to link biobank subjects to Taiwan’s own health insurance database and other longitudinal registries, which, similar to the Nordic countries, provide population-wide coverage of lifelong health information and events of nearly all of its citizens.

In companion with the Global Biobank Meta-Analysis Initiative (GBMI) flagship paper (Zhou et al., 2021), we provide here a detailed description of the TWB resource for East Asian genetics research and beyond. Different from previous publications on TWB that focused on specific aspects or analyses of the TWB data, we aim at giving an overview of TWB from cohort design, phenotype availability, genomic data generation, sample characteristics, genetic discoveries to date, and finally to its data access and sharing policy.

### Overview of the Taiwan Biobank (TWB)

The Taiwan Biobank (TWB) is a government-supported, prospective cohort study with a wide range of phenotypic measurements and genomic data collected on the Taiwanese population [https://www.twbiobank.org.tw/new_web/index.php].

Recruitment of TWB includes a community-based arm and a hospital-based collection. Commenced in 2012 with a target sample size of 200,000 individuals, the community-based arm has been enrolling men and women aged 30-70 with no prior diagnosis of cancer from more than 30 recruitment sites across Taiwan, distributed based on population density of different counties and cities. At recruitment, participants provided a written informed consent and had their baseline data collected through questionnaires, physical examination, and blood and urine tests (**Figure 1A; Table S1**). Repeated measurements of these phenotypes are planned to be implemented every 2 to 4 years. During the follow-up visits, participants would additionally undergo medical imaging examinations such as ultrasound and electrocardiogram scans. As of August 2021, a total of 151,406 volunteers have joined the biobank, among which 37,508 have completed the first round of follow-up. Whole-genome genotypes were measured for all participants, with a few other data types also available for selected subsets of the cohort, including whole-genome sequencing, DNA methylation, HLA typing, and blood metabolome (**Table S2**). The community-based data has been released to the scientific community on a regular basis in a de-identified format. The current cohort comprises more females than males (64% vs. 36%) and a similar number of participants across each 10-year age strata. 75% of the participants had a high school degree or higher at baseline (**Figure 1B**). Nearly all individuals are of Han Chinese descent (**Figure 2A**), reflective of the ancestry composition in Taiwan (Executive Yuan, Taiwan, R. O. C., 2016). The hospital-based arm, on the other hand, is a targeted collection starting in 2016 of patients affected with the most common chronic diseases in Taiwan and has not been made available to the public. We refer to TWB as the community-based cohort hereafter in this article.

**Figure 1.**
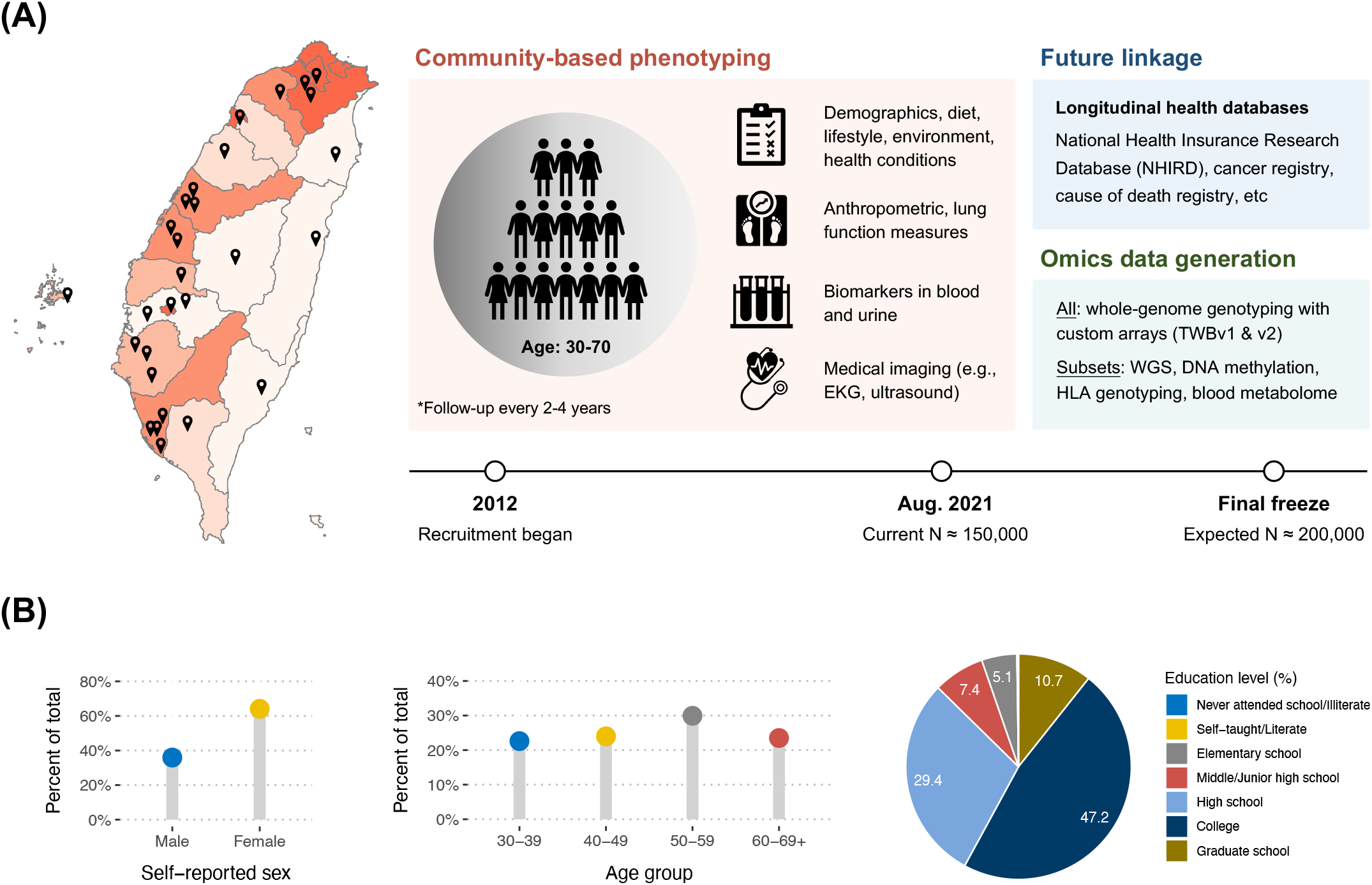
Design and demographics of the Taiwan Biobank (TWB) community-based cohort. **(A)** The community-based arm of TWB enrolled men and women aged 30-70 across recruitment centers in Taiwan that accounts for population density, with every county/city having at least one recruitment site (indicated by the location marks; a darker color represents a higher population density). Phenotypes were collected at baseline through a structured interview, physical examination, and blood/urine tests for each participant, with repeated measurements taken at a 2 to 4-year interval. As of August 2021, 37,508 individuals have finished the first round of follow-up among all 150,000 participants. Multi-omics data were generated for all or subsets of the participants, including array genotyping, whole-genome sequencing, HLA typing, DNA methylation, and blood metabolome (**Table S2**). A linkage to the NHIRD and other health-related registries is underway to provide phenotypic information in addition to self-report for the TWB. Enrollment is expected to continue until reaching the target of 200,000 volunteers. **(B)** The distribution of age, sex, and education level of study participants at baseline. More female than male participants were enrolled in TWB. Age of the participants was evenly distributed across every 10-year age bracket. People with a college degree accounted for the largest proportion in the current cohort.

**Figure 2.**
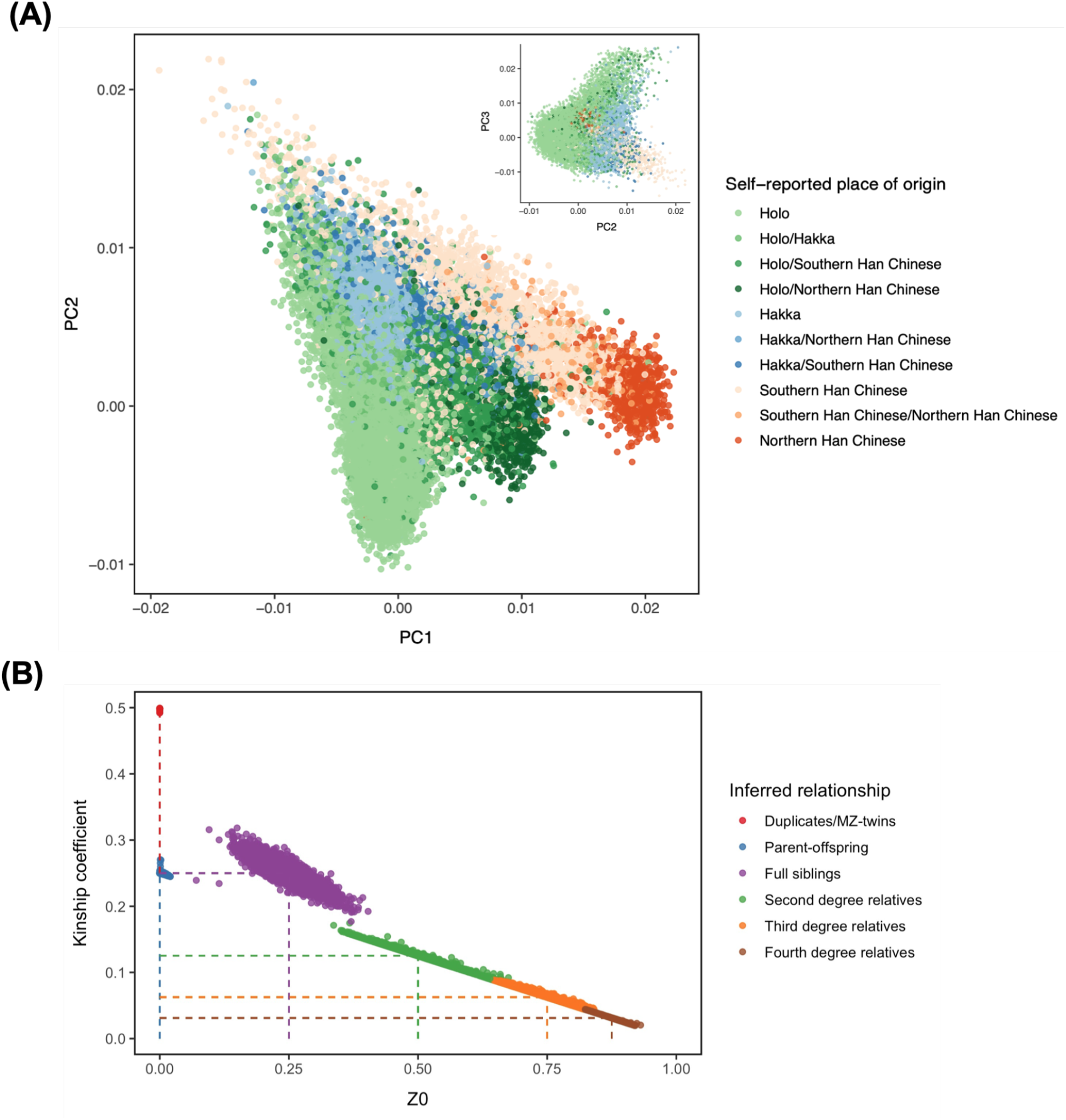
Population structure and familial relationship within TWB. **(A)** PCA on the QC’ed genotype data revealed a homogenous population structure of primarily Han Chinese descent among the TWB participants, which can be further divided into three distinct subgroups representing different geographic and ancestral origins (Holo, Hakka, and Mainlanders). Mainlanders were roughly separated into Southern and Northern Chinese for visualization (details in **Table S4**). Participants with the same paternal and maternal place of ancestral origin were assigned into one single subgroup; those with mixed origins were assigned with A/B labels. Projection of TWB onto 1KG data showed tight clustering with the EAS superpopulation as well as the two Han Chinese populations (**Figure S1**). Shown here are results from batch 2 of 66K individuals. PC plots for batch 1, while not shown, were nearly identical to the batch 2 results. **(B)** Kinship estimation of TWB participants showed a non-trivial number of relatedness, including over 25,000 pairs who are third-degree relatives or closer across batch 1 and batch 2 (**Table S5**). Plotted here are pairs of individuals in batch 2 with a kinship coefficient > 0.02 (y-axis; within fourth-degree relatedness), estimated using KING (Manichaikul et al., 2010), against the proportion of loci with 0 allele shared by descent (Z0; x-axis). Dashed lines indicate the midpoints of cutoffs commonly used for determining the pairwise familial relationship based on kinship coefficients (Duplicates/MZ twins: kinship > 0.353; Parent–offspring: 0.177 < kinship < 0.353 & Z0 < 0.05; Full siblings: 0.177 < kinship < 0.353 & Z0 > 0.05; Second-degree relatives: 0.088 < kinship < 0.177; Third-degree relatives: 0.044 < kinship < 0.088; Fourth-degree relatives: kinship < 0.044). A substantial number of TWB participants were found to be fourth-degree relatives, among whom a random subset of 10,000 pairs were included for demonstration in the figure.

### Phenotype sources and harmonization

The TWB primarily collected phenotypic data through a self-reported mechanism. Specifically, upon joining the biobank, participants would undergo a physical examination and a structured interview with a well-trained researcher to report demographics, lifestyle behaviors, environmental exposures, dietary habits, family history, and health-related information in a questionnaire. They also provided blood and urine specimens, which were used for laboratory assays of biomarkers and omics data generation. In particular, a total of 21 biomarkers were measured in biobank participants, indicative of a range of hematological, metabolic, kidney, and liver functions (**Table S3**). At each follow-up visit, a repeated assessment was carried out for phenotypes measured at baseline. Additionally, selected medical tests were conducted to generate medical images such as abdominal ultrasound, bone mass density, and electrocardiograms for assessing relevant health conditions (**Figure 1A**). A total of more than 1,000 phenotypes have been measured (**Table S1**) with selected sample characteristics shown in **Table 1**.

**Table 1.** Sample characteristics of TWB participants at baseline.

| Characteristic | Samples with available genotype data<sup>1</sup> |  |  | Analytical sample after QC<sup>2</sup> |  |
| --- | --- | --- | --- | --- | --- |
|  | All | Males | Females | Batch 1 (TWBv1) | Batch 2 (TWBv2) |
| Number of individuals | 108,955 | 39,410 | 69,545 | 27,033 | 65,582 |
| Age (mean ± SD) | 49.9 ± 10.9 | 49.9 ± 11.3 | 49.9 ± 10.7 | 48.8 ± 11.1 | 50.5 ± 10.7 |
| Sex (% women) | 63.8% | - | - | 50.0% | 68.8% |
| BMI (kg/m<sup>2</sup>; mean ± SD) | 24.2 ± 3.8 | 25.4 ± 3.5 | 23.6 ± 3.7 | 24.3 ± 3.7 | 24.2 ± 3.8 |
| Smoking experience (N; %) |  |  |  |  |  |
| Never or rarely | 87,742 (81%) | 21,934 (56%) | 65,808 (95%) | 20,555 (76%) | 55,190 (84%) |
| Ex-smoker | 10,993 (10%) | 9,282 (24%) | 1,711 (2.5%) | 3,272 (12%) | 6,191 (9.4%) |
| Current smoker | 10,218 (9.4%) | 8,192 (21%) | 2,026 (2.9%) | 3,194 (12%) | 5,623 (8.6%) |
| Alcohol consumption (N; %) |  |  |  |  |  |
| Never or rarely | 99,711 (92%) | 32,071 (81%) | 67,640 (97%) | 24,298 (90%) | 61,870 (94%) |
| Ex-drinker | 2,790 (2.6%) | 2,208 (5.6%) | 582 (0.8%) | 788 (2.9%) | 1,578 (2.4%) |
| Current drinker | 6,395 (5.9%) | 5,106 (13.0%) | 1,289 (1.9%) | 1,913 (7.1%) | 3,556 (5.4%) |
| **Quantitative measurements (mean ± SD)** |  |  |  |  |  |
| SBP (mmHg) | 119.3 ± 17.9 | 125.0 ± 16.5 | 116.1 ± 17.8 | 118.2 ± 17.4 | 119.7 ± 18.1 |
| LDL cholesterol (mg/dL) | 120.8 ± 31.7 | 121.7 ± 31.4 | 120.4 ± 31.8 | 120.9 ± 31.7 | 120.8 ± 31.6 |
| FEV1.0/FVC (%) | 79.8 ± 17.5 | 79.7 ± 17.4 | 79.9 ± 17.5 | 73.1 ± 18.8 | 81.1 ± 17.1 |
| Fasting glucose (mg/dL) | 95.9 ± 20.6 | 99.4 ± 23.4 | 93.9 ± 18.5 | 96.3 ± 21.0 | 95.7 ± 20.5 |
| Creatinine (mg/dL) | 0.72 ± 0.31 | 0.91 ± 0.37 | 0.62 ± 0.21 | 0.76 ± 0.36 | 0.71 ± 0.30 |
| Albumin (g/dL) | 4.52 ± 0.23 | 4.59 ± 0.24 | 4.48 ± 0.22 | 4.56 ± 0.24 | 4.50 ± 0.23 |
| **Most common self-reported diseases in TWB (N; %)** |  |  |  |  |  |
| Peptic ulcer | 15,926 (15%) | 6,548 (17%) | 9,378 (13%) | 3,923 (15%) | 9,897 (15%) |
| GERD | 14,682 (13%) | 5,054 (13%) | 9,628 (14%) | 3,306 (12%) | 9,145 (14%) |
| Hypertension | 13,432 (12%) | 6,667 (17%) | 6,765 (9.7%) | 3,297 (12%) | 8,445 (13%) |
| Drug medication | 10,243 (9.4%) | 2,934 (7.4%) | 7,309 (11%) | 2,403 (8.9%) | 6,485 (9.9%) |
| Hyperlipidemia | 8,019 (7.4%) | 3,615 (9.2%) | 4,404 (6.3%) | 1,884 (7.0%) | 5,020 (7.7%) |
| Kidney stone | 7,003 (6.4%) | 4,225 (11%) | 2,778 (4.0%) | 1,973 (7.3%) | 4,116 (6.3%) |

<sup>1</sup>TWB samples with available genome-wide genotyping data at the time of our analysis<sup>2</sup>The final analytical sample after batch-specific genotype QC; see details in Chen et al., 2021. The two batches correspond to the two different custom arrays (TWBv1 and v2) used in TWB. Batch-specific results are then combined via meta-analysis.

In addition to self-reported medical conditions, disease and health information of a participant can be obtained through linkage to the National Health Insurance Research Database (NHIRD) and >70 additional databases that cover specific health-related outcomes in the Taiwanese population, such as the Taiwan Cancer Registry and cause of death registry (Hsieh et al., 2019). Taiwan’s National Health Insurance (NHI) is a single-payer compulsory insurance program instituted in 1995 that provides accessible and affordable health care to all citizens in Taiwan (coverage >99.99%) (National Health Insurance Administration, Ministry of Health and Welfare, Taiwan, R. O. C., 2014). The NHIRD is constructed for research purposes from registration and claims data in the NHI, actively maintained since 2002, and contains an extensive list of sorted data files, such as registry for beneficiaries, drug prescription registry, inpatient claims, registry for medical facilities, and three embedded longitudinal health insurance databases (Hsieh et al., 2019). Through linking to the unique personal identification numbers assigned to each individual in the NHIRD, data available only in other registries and regional hospitals may also be acquired. For example, founded in 1979, the Taiwan Cancer Registry includes population-based collection of detailed cancer staging, treatment, and recurrence information for both in- and out-patients diagnosed with malignant neoplasms (Chiang et al., 2019). Cross-linkage between TWB and these health care databases currently only permits “on-site” analysis with a separate, study-specific Institutional Review Board (IRB) approval. Efforts are underway to integrate health-related records from across these databases with the TWB community-based phenotype collection. Genetic analysis of TWB that contributed to the GBMI effort focused on self-reported diseases at baseline.

### Genotype generation, quality control, and imputation

Based on blood samples collected upon recruitment, genome-wide genotype data was available for 108,955 participants at the time of our analysis. Two different customized arrays (TWBv1 and TWBv2) were used for genotyping based on the GRCh37/hg19 coordinates. The first subset of 27,719 participants were genotyped on the TWBv1 array, which was designed based on Thermo Fisher Axiom Genome-Wide CHB Array with customized contents in 2011, including ∼650,000 markers. The remaining 81,236 participants were genotyped using the TWBv2 array, designed by Thermo Fisher Scientific Inc. in 2017 based on whole-genome sequencing data from 946 TWB samples with customized contents enriched for rare coding risk alleles, including ∼690,000 markers (Juang et al., 2021; Wei et al., 2021). The two arrays share only ∼100,000 markers.

For the TWB-GBMI analysis demonstrated in the flagship paper of this special issue (Zhou et al., 2021), we included all individuals genotyped on TWBv1 (batch 1) and ∼80% of those genotyped on TWBv2 (batch 2) as the discovery sample for genome-wide association studies (GWAS) (Chen et al., 2021; **Table 1**).

Considering differences in marker content between the two genotyping arrays, our quality control (QC) procedures were performed for each batch separately using custom scripts adapted from [https://github.com/Annefeng/PBK-QC-pipeline]. For pre-imputation QC, we first removed variants with call rate < 0.98, samples with call rate < 0.98, as well as variants that were monomorphic, duplicated, or not confidently mapped to a genomic position. Next, we inferred genetic ancestry of TWB participants using 1000 Genomes (1KG) phase 3 samples (The 1000 Genomes Project Consortium et al., 2015) as the population reference panel. Specifically, we merged the TWB data with the 1KG data and performed principal component analysis (PCA) based on high-quality, common (minor allele frequency, or MAF, > 5%), bi-allelic single nucleotide polymorphisms (SNPs) in approximate linkage equilibrium (LD). Using the 1KG data as the training set, we then fit a Random Forest classifier with the first 6 principal components (PCs) to assign ancestry of TWB participants at a prediction probability > 0.8 into each of the five 1KG super-populations: European (EUR), African (AFR), Admixed/Latino American (AMR), Eat Asian (EAS), and South Asian (SAS). Focusing on the predicted EAS ancestral group that contained the majority of the TWB participants, we further removed samples whose self-reported and genetic sex did not match and those with outlying values of autosomal heterozygosity rate. To identify any residual population outliers, we performed three rounds of within-EAS PCA, which sequentially removed samples with any of the top 10 PCs deviating from the mean by more than 6 standard deviations. Lastly, we filtered out variants with call rate < 0.98 and Hardy-Weinberg equilibrium (HWE) test p-value < 1×10^−10^.

Based on this homogenous EAS sample, we computed the final in-sample PCs to be used in subsequent analyses, and performed pre-phasing using Eagle v2.4 (Loh et al., 2016) and genotype imputation using Minimac4 (Fuchsberger et al., 2015) with 1KG phase3 EAS dataset as the reference panel. At INFO > 0.6, MAF distribution in the imputed data showed that each batch contained ∼10% very rare variants (MAF < 0.001), ∼15% rare variants (0.001 < MAF < 0.01), ∼15% low-frequency variants (0.01 < MAF < 0.05), and ∼60% common variants (0.05 < MAF <0.5) (Figure S1). After imputation, we retained variants with an imputation quality INFO score > 0.6 and MAF > 0.5%. The final analytic sample consisted of a total of 8,190,806 variants in 27,033 individuals for batch 1, and 8,156,315 variants in 65,582 individuals for batch 2, respectively.

### Population structure and familial relatedness

As revealed from self-report and a series of PCA, the population structure of TWB is highly homogenous, comprising Taiwanese individuals of predominantly Han Chinese descent (>99%; **Figures 2A, S2A & S2B**), largely consistent with the racial/ethnic demographic of Taiwan from the national census (Executive Yuan, Taiwan, R. O. C., 2016). Based on the QC’ed genotype dataset of TWB, the first two PCs distinguished individuals from various sub-continental geographic origins, including a majority (∼78%) who self-identified as Holo (or Hoklo) or Hakka— the two largest Han Chinese groups in Taiwan who arrived from Southeast China during the last few centuries—as well as those who migrated to Taiwan after 1949 (“Mainlanders”) and their descendants with a paternal/maternal ancestral origin that can be traced back to different provinces across Mainland China (∼6-7%; **Figure 2A; Table S4**). Interestingly, while Holo and Hakka people both originated from the Southeast coast of China in adjacent regions—mainly the Fujian and Guangdong provinces—PCA suggested a differentiation in genetic variation between the two ethnic groups, likely attributable to physical boundaries separating the two regions. Among the Mainlanders, as well as those who have mixed ancestral origins between Holo, Hakka, and the Mainlanders, a North-South gradient of genetic variation was also observed, which has been previously shown to correspond with the migration history of the Han Chinese population (Chen et al., 2009). Further PCs formed a single ball-shaped cluster when plotted against each other, showing no additional signs of sub-structure (**Figure S2C**).

Using genetic data to identify familial relationships, our kinship estimation among TWB participants inferred more than 25,000 related pairs within third-degree or closer, including 9 pairs of duplicates or MZ twins, 4,154 parent-offspring pairs, 5,079 sibling pairs, 2,756 pairs of second-degree relatives, and >10,000 pairs of third-degree relatives across the two batches (**Table S5**; batch 2 results in **Figure 2B**). The non-trivial number of pairwise relatedness could be due to recruitment that took place in local communities, whereby family members might live in proximity and were more likely to be recruited together or invite one another to participate when informed of the TWB activity.

### Genetic association analysis

Considering the substantial genetic relatedness in TWB and to maximize power, we performed GWAS analysis using REGENIE (Mbatchou et al., 2021), a computationally efficient two-step whole-genome regression method for large-scale genomics analysis that accounts for sample relatedness and population structure and offers superior statistical power over analysis restricting to unrelated individuals. In brief, we computed in step 1 a leave-one-chromosome-out (LOCO) polygenic score combining estimates across blocks of SNPs using two levels of Ridge regression. At step 2 we performed association testing using logistic regression model score test that included the LOCO predictors as an offset in addition to a set of covariates (age, age2, sex, age x sex, age2 x sex, and the top 20 PCs). Association test statistics from batch 1 and batch 2 were then meta-analyzed using the inverse-variance-weighted fixed-effect model based on ∼7.7M overlapping SNPs between the two batches. Assessment of our TWB GWAS results of dozens of quantitative traits and self-reported diseases suggested no inflation in association statistics due to population stratification (Chen et al., 2021).

### Unique attributes and weaknesses of the biobank

Aside from phenotypes commonly shared across different biobanks, TWB measured lifestyle traits and environmental exposures that are more specific to the Taiwanese population as part of East and Southeast Asia. For example, included in the questionnaires were questions regarding exposure to different types of incense burning, such as mosquito repellent coils and joss sticks used in religious rituals, and betel nut chewing, a cultural tradition that has been associated with adverse health risk (Gupta and Warnakulasuriya, 2002). In addition, TWB employed a 44-item Body Constitution Questionnaire (Lin et al., 2012) that, according to traditional Chinese medicine theories, classifies individuals into one or more body constitution types, based upon which one’s susceptibility to specific symptoms or diseases can be inferred to make personalized recommendations for promoting health-conscious lifestyle.

One noticeable issue of TWB is that data might not be population-representative from community-based sampling, with visibly more women than men and no children in the cohort. Based on self-report alone, several common diseases also showed an in-sample prevalence lower than that in the general adult population (e.g., hypertension, 12.5% in TWB vs. 26% reported from the Ministry of Health and Welfare in Taiwan for adults aged 20 and above; **Table S4**), which may reflect biases from self-report or similar to what was observed in the UK Biobank of a healthy volunteer bias (Fry et al., 2017). On the other hand, with future linkage to the NHIRD and other registries, TWB will provide a useful resource for studying genetic and environmental liability to a broad spectrum of diseases and longitudinal outcomes beyond self-report in the Han Chinese population, including those traditionally found to be more prevalent in Taiwan and its neighboring areas, such as nasopharyngeal carcinoma and liver diseases (Chang and Adami, 2006; Sarin et al., 2020).

### Genetic discoveries based on TWB

Several works utilizing TWB data have demonstrated its value in human genetic studies and expanded population diversity in genomic research in East Asia. Based on the initial release of 10,000 individuals, an earlier investigation provided a deeper dive into the population structure of TWB, showing that, in addition to Northern and Southern Han Chinese, there was a third cluster genetically similar to the Southern Han Chinese but with an extended haplotype in the major histocompatibility complex (MHC) region, potentially reflecting a more recent evolutionary event (Chen et al., 2016). Leveraging different kinds of physical activity measures in TWB, researchers have found that several exercises, such as regular jogging and mountain climbing, can attenuate the genetic influences on obesity and BMI, suggesting converging evidence with previous findings from EUR-centered genetic studies (Lin et al., 2019). Measured in a subset of TWB participants, the whole-genome sequencing data of at least 30X coverage among 1,445 individuals from TWB has been demonstrated to outperform the 1KG EAS samples as an imputation panel for Han Chinese-ancestry individuals, providing a moderate yet consistent improvement (Wei et al., 2021). Exploring this Han-Chinese-specific reference panel also led to the identification of a great number of population-specific rare variants, many of which have not been seen in existing large allele frequency databases (e.g., gnomAD (Karczewski et al., 2020)), which has the potential to facilitate discovery of rare disease-causing variants, and, combined with the available information on common genetic variation from TWB array data, to improve genomic prediction for disease (e.g., hypertension) (Juang et al., 2021). More recently, we examined the genetic architecture of 36 quantitative traits, including anthropometric traits and biomarkers, across TWB, Biobank Japan, and UK Biobank, in which we identified TWB-specific novel genetic loci, fine-mapped those to putative causal variants, showed that genetic architecture of these traits was largely consistent within EAS as well as between EAS and EUR populations, and suggested the utility of biomarker GWAS in polygenic risk prediction for complex diseases in cross-population, multi-trait settings (Chen et al., 2021).

### Data access and sharing of the TWB data

All data including individual genotypes and phenotypes of TWB participants is available upon application for research purposes. A detailed description of TWB data availability and the application process can be found at https://taiwanview.twbiobank.org.tw/data_appl. In brief, investigators who are interested in obtaining the TWB data would need to submit an application that includes a detailed research proposal and an institutional review board (IRB) approval from the applicant’s home institute to the TWB Data Release Group (contact email). The application will go through scientific and ethical reviews by external experts in the relevant scientific fields and the Ethics and Governance Committee (EGC) of TWB. Once approved, researchers will be able to obtain the data for the approved research projects during the approved time period. For researchers who are interested in applying but reside outside of Taiwan, or any cross-country collaborations, an additional international data transfer agreement needs to be filed to the Ministry of Health and Welfare of Taiwan to enable sharing of the TWB individual-level data or any of its derivatives.

## Conclusions

The Taiwan Biobank (TWB) is transforming biomedical research into the genetic basis of health and disease in the Taiwanese population. Here we present an overview of the TWB resource (N∼150k in 2021), illustrate the population structure and familial relationship within TWB, and highlight unique aspects and current genetic findings from TWB. In the next few years, TWB will continue to grow in sample size, accumulate longitudinal and repeated measures, and generate molecular phenotypes on the enrolled subjects. With the endeavor to link subjects to the NHIRD, which covers almost all citizens of Taiwan since 1995, and other registries, TWB will develop into an invaluable resource integrating rich phenotypes and multi-omics data across a long timeframe, delivering its mission of improving public health in Taiwan. Being one of the largest biobanks in East Asia, TWB will also contribute to the insights of human complex disorders and traits in world populations through integrative and comparative study with other global biobanks, as demonstrated by a series of studies in this special issue and beyond (Chen et al., 2021; Zhou et al., 2021).

## Data Availability

All data including individual genotypes and phenotypes of Taiwan Biobank (TWB) participants is available upon application for research purposes. A detailed description of TWB data availability and the application process can be found at https://taiwanview.twbiobank.org.tw/data_appl. In brief, investigators who are interested in obtaining the TWB data would need to submit an application that includes a detailed research proposal and an institutional review board (IRB) approval from the applicant's home institute to the TWB Data Release Group (contact). The application will go through scientific and ethical reviews by external experts in the relevant scientific fields and the Ethics and Governance Committee (EGC) of TWB. Once approved, researchers will be able to obtain the data for the approved research projects during the approved time period. For researchers who are interested in applying but reside outside of Taiwan, or any cross-country collaborations, an additional international data transfer agreement needs to be filed to the Ministry of Health and Welfare of Taiwan to enable sharing of the TWB individual-level data or any of its derivatives.

## Acknowledgement

This research has been conducted using the Taiwan Biobank resource. We thank all the participants and investigators of the Taiwan Biobank. We thank the National Center for Genome Medicine of Taiwan for the technical support in genotyping. We thank the National Core Facility for Biopharmaceuticals (NCFB, MOST 106-2319-B-492-002) and National Center for High-performance Computing (NCHC) of National Applied Research Laboratories (NARLabs) of Taiwan for providing computational and storage resources. Y.F.L. is supported by the National Health Research Institutes (NP-109-PP-09; NP-110-PP-09) and the Ministry of Science and Technology (109-2314-B-400-017; 110-2314-B-400 -028 -MY3) of Taiwan. T.G. is supported by NIA K99/R00AG054573. H.H. acknowledges support from NIDDK K01DK114379, NIMH U01MH109539, Brain & Behavior Research Foundation Young Investigator Grant, the Zhengxu and Ying He Foundation, and the Stanley Center for Psychiatric Research. Y.A.F. and P.H.K. are supported by the “National Taiwan University Higher Education Sprout Project (NTU-110L8810)” within the framework of the Higher Education Sprout Project by the Ministry of Education (MOE) in Taiwan. YH.H. acknowledges support from NIAMS R01AR072199.

## Ethics

The access to and use of the Taiwan Biobank data in the present work was approved by the Ethics and Governance Council (EGC) of Taiwan Biobank (approval number: TWBR10907-05) and the Institutional Review Board (IRB) of National Health Research Institutes, Taiwan (approval number: EC1090402-E). The data collection of Taiwan Biobank was approved by the Ethics and Governance Council (EGC) of Taiwan Biobank and the Department of Health and Welfare, Taiwan (Wei-Shu-I-Tzu NO.1010267471). Taiwan Biobank obtained informed consent from all participants for research use of the collected data and samples.

## Supplemental Text and Figures

**Table S1.** Summary of phenotypes collected in the TWB community-based cohort at baseline and follow-up visits through questionnaires, physical examinations, and blood and urine tests.

| Category | Phenotypes |
| --- | --- |
| **Demographics** | Age, sex, date of birth, education, marriage, job experience, income, place of paternal/maternal ancestral origin (Hoklo/Hakka/Aboriginal/Waishengren), etc. |
| **Behavioral** | Alcohol consumption, smoking behavior, betel nut usage, physical activity, weight change, drug use, sleep pattern, etc. |
| **Environmental** | Cooking habits, incense exposure, water supply, etc. |
| **Diet** | Food choice, tea/coffee consumption, vegetarian diet, mealtime, supplement use, etc. |
| **Anthropometric** | Body height, body weight, waist/hip circumference, body fat rate, etc. |
| **Medical images** (starting from the 1<sup>st</sup> follow-up) | Abdominal ultrasonography, bone mass density, electrocardiography, vertebral artery ultrasound, and thyroid gland ultrasound |
| **Health condition-related** | (including family history and month/year of diagnosis when applicable) |
| *Respiratory* | Lung function (e.g., FVC, FEV1), asthma, COPD, emphysema/bronchitis, etc. |
| *Cardiovascular* | Systolic/diastolic pressure, coronary artery disease, hypertension, etc. |
| *Endocrine/Metabolic* | Diabetes yes/no, type of diabetes, etc |
| *Digestive* | Peptic ulcer, GERD, irritable bowel disease, liver/gall stone, etc. |
| *Genitourinary* | Kidney stone, renal failure, female-specific conditions (e.g., menarche, menopause, myoma, ovarian cyst, endometriosis), etc. |
| *Musculoskeletal* | Bone density, arthritis, gout, osteoporosis, etc. |
| *Psychological* | Psychosis, depression, schizophrenia, OCD, alcoholism, mental health questionnaires (PHQ-4 and MMSE), etc. |
| *Neurological* | Epilepsy, Parkinson's disease, dementia, hemicrania, multiple sclerosis, etc. |
| *Eye* | Cataract, glaucoma, xerophthalmia, floaters, color blindness, etc. |
| *Pain* | Headache, sciatica, joint pain, neck ache, etc. |
| *Carcinoma in situ* | Liver/lung/breast/gastric/colorectal/nasopharyngeal/prostate/other |
| *Drug allergy* | Allergy yes/no, type of medications, etc. |
| *Traditional Chinese medicine constitution* | Frequent phlegm, cold or warm hands/feet, fatigue, ringing in ears, shortness of breath, dry eyes, dizziness, etc. |
| **Biomarkers** | RBC, WBC, platelet, HbA1c, HBsAg, triglyceride, bilirubin, creatinine, etc. |

**Table S2.** Summary of available multi-omics datasets within TWB (as of August 2021)

| **Data type** | **Sample size** | **Note** |
| --- | --- | --- |
| Whole-genome genotyping | All participants (150,000) | Customized chips: TWB v1.0 of 650K genetic markers and TWB 2.0 of 690K genetic markers |
| Whole-genome sequencing | 2,000 | Coverage 30X |
| DNA methylation | 1,100 | Illumina Infinium MethylationEPIC 850K BeadChip |
| HLA typing | 2,100 | Class I: HLA-A, HLA-B, HLA-C; class II: HLA-DPA1, HLA-DPB1, HLA-DQA1, HLA-DQB1, HLA-DRB1, HLA-DRB345 |
| Blood metabolome | 760 | Profiled by NMR spectroscopy |

**Table S3.**
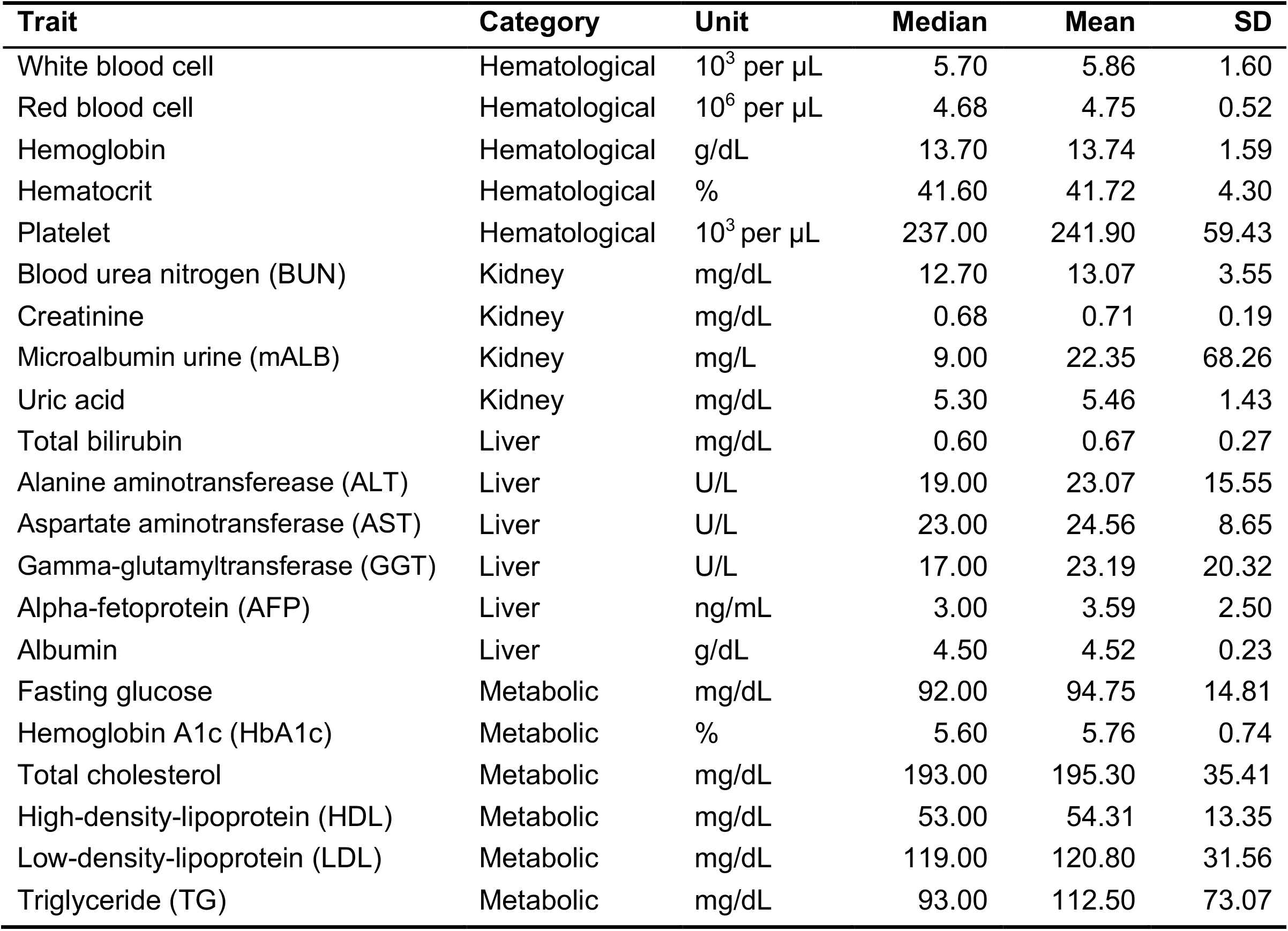
21 biomarker measurements at baseline in the current analytical sample of TWB (N=92,615)

**Table S4.** Classification of self-reported ancestry of TWB participants (N=92,615)

| **Self-reported place of origin** | **Batch1** | **Batch2** | **All** |
| --- | --- | --- | --- |
| Holo | 17,844 | 44,062 | 61,906 (66.6%) |
| Holo/Hakka | 1,519 | 4,015 | 5,534 (6.0%) |
| Holo/Southern Han Chinese | 1,572 | 3,520 | 5,092 (5.5%) |
| Holo/Northern Han Chinese | 479 | 1,106 | 1,585 (1.7%) |
| Hakka | 2,715 | 8,005 | 10,720 (11.5%) |
| Hakka/Northern Han Chinese | 89 | 275 | 364 (0.4%) |
| Hakka/Southern Han Chinese | 360 | 930 | 1,290 (1.4%) |
| Southern Han Chinese* | 1,613 | 3,199 | 4,812 (5.2%) |
| Southern/Northern Han Chinese* | 143 | 297 | 440 (0.5%) |
| Northern Han Chinese* | 363 | 781 | 1,144 (1.2%) |
| **Total N** | **27,033** | **65,582** | **92,615** |

\*The “Northern Han Chinese” group included those who reported a paternal or maternal ancestry of origin from the following 22 provinces in China prior to 1949: Shanxi, Shandong, Gansu, Andong, Hejiang, Jilin, Songjiang, Hebei, Henan, Qinghai, Shaanxi, Heilongjiang, Suiyuan, Xinjiang, Chahar, Nenjiang, Ningxia, Mongolia, Jehol, Liaobei, Liaoning, and Xing'an; alternatively, the “Southern Han Chinese” category was based on self-reported origin in the following 15 provinces: Sichuan, Jiangsu, Jiangxi, Anhui, Xikang, Tibet, Zhejiang, Hainan, Hubei, Hunan, Guizhou, Yunnan, Fujian, Guangdong, and Guangxi.

**Table S5.** Number of related pairs within third-degree relatedness inferred by kinship estimation in TWB (N=92,615)

| **Inferred familial relationships** | **Batch1** | **Batch 2** |
| --- | --- | --- |
| Duplicates/MZ twins | 7 | 22 |
| Parent-offspring | 827 | 3,327 |
| Full siblings | 952 | 4,127 |
| 2<sup>nd</sup> degree relatives | 428 | 2,337 |
| 3<sup>rd</sup> degree relatives | 1,128 | 12,347 |

**Table S4.** Comparison of prevalence of selected self-reported common diseases in TWB (N=92,615) and the general adult population in Taiwan.

| **Self-reported disease in TWB** | **Within-TWB prevalence** | **Reported population prev.** | **Reference or PMID** |
| --- | --- | --- | --- |
| Hypertension | 12.5% | 26% | MOHW* statistics 2013-2019, age ≥ 20; 31816161 |
| Hyperlipidemia | 7.34% | 22% | MOHW statistics 2013-2019, age ≥ 20 |
| Diabetes | 5.11% | 9-12%; | MOHW statistics 2013-2019, age ≥ 20 |
| Gout | 3.94% | 4-6% | 25612613; 24265412 |
| Depression | 3.56% | 1.2% | 22051196 |
| Asthma | 3.43% | 11% | MOHW statistics 2000-2011, adults |
| Emphysema or bronchitis (COPD) | 1.21% | 6-10% | 25937817; 26648708 |
| Apoplexia (Stroke) | 0.62% | 1.6% | 2749845 |

\*MOHW: Ministry of Health and Welfare

**Figure S1.**
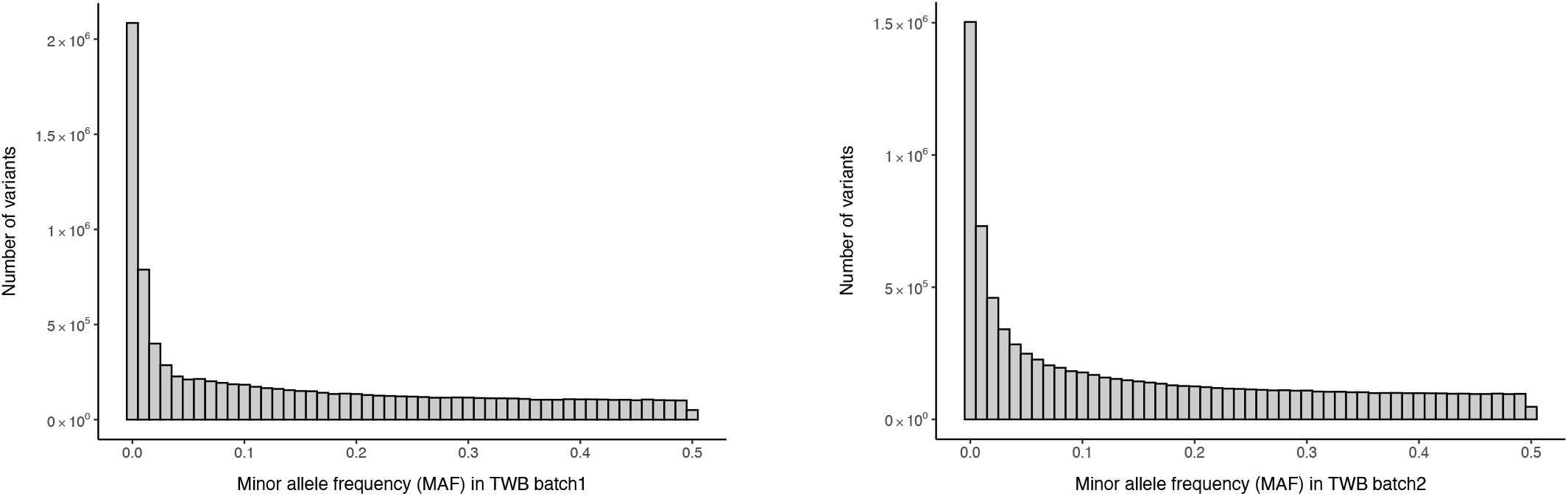
MAF distribution in the TWB imputed dataset (N=92,615) at INFO > 0.6. The plots show the MAF distribution of genetic markers with an imputation INFO score > 0.6 based on all individuals in batch 1 (left) and batch 2 (right), separately. Both batch 1 and batch 2 contained <10% “very rare” variants (MAF < 0.001), ∼15% “rare” variants (0.001 < MAF < 0.01), ∼15% “low frequency” variants (0.01 < MAF < 0.05), and ∼60% “common” variants (0.05 < MAF <0.5).

**Figure S2.**
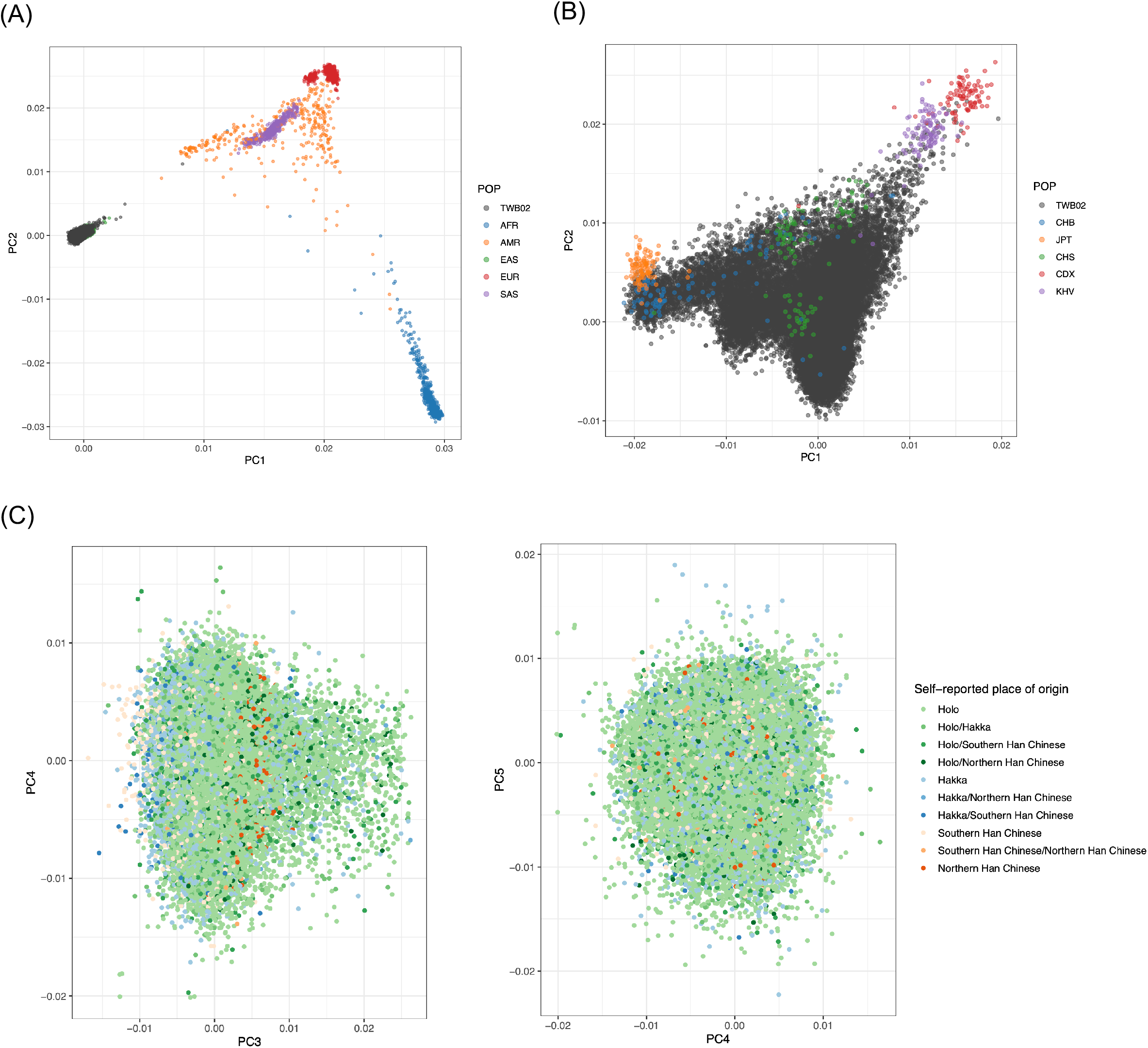
Population structure of TWB based on batch 2 of 66K individuals. (A) TWB participants clustered with the 1000 genomes EAS (East Asian) superpopulation (B) TWB participants clustered with the 1000 genomes CHB (Han Chinese in Beijing) and CHS (Han Chinese from Southern China) populations (C) PC plots of PC3 and later showed a homogenous cluster with no obvious sub-structure. A corresponding figure of PC1 vs. PC2 and PC2 vs. PC3 is shown in **Figure 2A**. Population structure among batch1 participants, while not shown, is nearly identical to that observed in batch 2.

